# Critical care workers have lower seroprevalence of SARS-CoV-2 IgG compared with non-patient facing staff in first wave of COVID19

**DOI:** 10.1101/2020.11.12.20145318

**Authors:** H.E. Baxendale, D. Wells, J. Gronlund, A. Nadesalingam, M. Paloniemi, G. Carnell, P. Tonks, L. Ceron-Gutierrez, S. Ebrahimi, A. Sayer, J.A.G. Briggs, X. Xiong, J.A. Nathan, G.L. Grice, L.C. James, J. Luptak, S. Pai, J.L. Heeney, R. Doffinger

## Abstract

With the first 2020 surge of the COVID-19 pandemic, many health care workers (HCW) were re-deployed to critical care environments to support intensive care teams to look after high numbers of patients with severe COVID-19. There was considerable anxiety of increased risk of COVID19 for staff working in these environments.

Using a multiplex platform to assess serum IgG responses to SARS-CoV-2 N, S and RBD proteins, and detailed symptom reporting, we screened over 500 HCW (25% of the total workforce) in a quaternary level hospital to explore the relationship between workplace and evidence of exposure to SARS-CoV-2.

Whilst 45% of the cohort reported symptoms that they consider may have represented COVID-19, overall seroprevalence was 14% with anosmia and fever being the most discriminating symptoms for seropositive status. There was a significant difference in seropositive status between staff working in clinical and non-clinical roles (9% patient facing critical care, 15% patient facing non-critical care, 22% nonpatient facing). In the seropositive cohort, symptom severity increased with age for men and not for women. In contrast, there was no relationship between symptom severity and age or sex in the seronegative cohort reporting possible COVID-19 symptoms. Of the 12 staff screened PCR positive (10 symptomatic), 3 showed no evidence of seroconversion in convalescence.

**Conclusion:** The current approach to Personal Protective Equipment (PPE) appears highly effective in protecting staff from patient acquired infection in the critical care environment including protecting staff managing interhospital transfers of COVID-19 patients. The relationship between seroconversion and disease severity in different demographics warrants further investigation. Longitudinally paired virological and serological surveillance, with symptom reporting are urgently required to better understand the role of antibody in the outcome of HCW exposure during subsequent waves of COVID-19 in health care environments.

## Introduction

In December 2019, clusters of patients with novel pneumonia-like symptoms were identified in Wuhan, China (1). This disease is now known as COVID-19 and is caused by the SARS-CoV-2 Coronavirus. Since the initial cases, the virus has so far infected over 35 million people causing over 1,000,000 deaths globally (2). In response to this pandemic many countries have and continue to employ social distancing practices and many working from home has become the norm for desk-based employments.

There is limited opportunity for healthcare workers to comply with social distancing measures due to the need for patient contact. Their risk of infection may be especially high as patients with high viral load are more likely to have severe disease and thus be admitted to hospital for earlier treatment (3, 4). In addition, representation of staff from Black, Asian and Minority Ethnic groups (BAME) is disproportionately high within health care (5), noting that individuals identifying as BAME appear to be at increased risk of severe COVID-19 (6). As such, guidelines for managing exposure to SARS-CoV-2 is now modified to take account of ethnicity risk additional to previously acknowledged risk factors such as age, sex and co-morbidities(7).

In this study, we investigated the prevalence of COVID-19 symptoms and evidence of exposure to SARS-CoV-2 through serum antibody assessment in a UK regional critical care and extracorporal membrane oxygenation centre (ECMO), Royal Papworth Hospital NHS Trust (RPH), from April to June 2020. We stratified our results according to staff demographic and workplace. RPH is a UK hospital specialising in cardiothoracic medicine and critical care which has been involved in supporting the regional COVID-19 critical care response including retrievals of ventilated patients from across the region and establishing ECMO at retrieval hospitals prior to transfer. We used an in house developed Luminex multiplex serology assay to detect serum IgG to nucleocapsid (N), spike (S) and the spike protein receptor binding domain (RBD) SARS-CoV2 antigens to determine evidence of previous virus exposure.

This paper highlights: (1) the detailed methodology for the multiplex COVID-19 serological testing; (2) the relationship between SARS-CoV-2 N, S and RBD antibodies in staff in clinical and non-clinical roles; (3) the distribution of symptoms within seropositive and seronegative cases, and (4) the risk factors for more severe COVID19 based on intersecting categories of age group, ethnicity and sex.

## Methods

### Recruitment

Staff from Royal Papworth Hospital were recruited through staff email over the course of 2 months (20^th^ April 2020-10^th^ June 2020) as part of a prospective study to establish seroprevalence and immune correlates of protective immunity to SARS-CoV-2.

Following informed consent, staff were invited to complete a questionnaire to clarify whether they had swab PCR confirmed SARS-CoV-2 infection and whether they had experienced symptoms that they felt may have been consistent with COVID-19 since January 2020. They were asked to describe these symptoms in free text, when they occurred and how long they lasted. Additional information including age (in years), sex, ethnicity, job classification (patient facing/non-patient facing), and whether they had been exposed to COVID-19 patients over the prior 4 months, was also provided.

Symptom severity was classified according to WHO severity classification into asymptomatic, mild, moderate and severe disease (8). Staff with dyspnoea additional to cough and fever were classified as moderate and staff requiring oxygen were classified as severe disease severity. Symptoms reported associated with COVID-19 were then tabulated from the free text responses provided. Reported symptoms included: anosmia, cough, dyspnoea, fatigue, fever, gastrointestinal (GI) disturbance, headache, myalgia, pharyngitis, upper respiratory infection (URTI).

Blood was taken and serum isolated for analysis of IgG binding to SARS-CoV-2 Spike, Nucleocapsid and RBD proteins using a Luminex based multiplexed particle flow cytometry assay developed at Addenbrookes Hospital NHS Trust. The assay was validated on PCR confirmed positive COVID-19 patients and pre-COVID-19 healthy control serum samples and showed excellent sensitivity and specificity (Supp Fig 1a: ROC curve analysis).

500 staff members were recruited and serum binding to all three antigens was quantified for 498 samples.

### SARS-CoV-2 serology by multiplex particle-based flow cytometry (Luminex)

Recombinant SARS-CoV-2 N, S and RBD were covalently coupled to distinct carboxylated bead sets (Luminex; Netherlands) to form a 3-plex assay. The S protein construct used is S-R/PP as described in Xiong et al 2020 (6, 9). The RBD protein construct used is described by Stadlbauer et al (10).

Beads were first activated with 1-ethyl-3-[3-dimethylaminopropyl]carbodiimide hydrochloride (Thermo Fisher Scientific) in the presence of N-hydroxysuccinimide (Thermo Fisher Scientific), according to the manufacturer’s instructions, to form amine-reactive intermediates. The activated bead sets were incubated with the corresponding proteins at a concentration of 50 μg/ml in the reaction mixture for 3 h at room temperature on a rotator. Beads were washed and stored in a blocking buffer (10 mM PBS, 1% BSA, 0.05% NaN3).

The N-, S- and RBD-coupled bead sets were incubated with proband sera at a 1/100 dilution for 1 h in 96-well filter plates (MultiScreen HTS; Millipore) at room temperature in the dark on a horizontal shaker. Fluids were aspirated with a vacuum manifold and beads were washed three times with 10 mM PBS/0.05% Tween 20. Beads were incubated for 30 min with a PE-labeled anti–human IgG-Fc antibody (Leinco/Biotrend), washed as described above, and resuspended in 100 μl PBS/Tween. They were then analyzed on a Luminex analyzer (Luminex / R&D Systems) using Exponent Software V31. Specific binding was reported as mean fluorescence intensities (MFI).

### Statistical methods

#### Serostatus classification

We trained a linear support vector machine (SVM) in scikit-learn (11) to classify samples as seropositive or seronegative based on Luminex readings of SARS-CoV-2 N, S, and S-RBD (Figure S1b: dataset). Our training set was 101 hospitalised patients with confirmed COVID-19 and 126 pre-pandemic control samples. Luminex readings were centred and scaled by the mean and standard deviation of the training set. To assess the accuracy of our SVM we used 5-fold cross validation, preserving the ratio of seropositive to seronegative cases. Then a model was trained on the whole training set and used to classify Papworth staff cohort, after centering and scaling according to the training set mean and standard deviation. Receiver operator analysis (ROC) was performed using GraphPad-Prism8 software.

### Risk of infection

We estimated the risk of infection, measured by serostatus, amongst Royal Papworth Hospital staff based on their area of work. Staff were classified as critical-care patient facing, non-critical-care patient facing, and non-patient facing. Binomial logistic regression was used to estimate infection risk using area of work as a predictor variable. The significance of the two patient facing areas was assessed by Wald Z-tests in contrast to the non-patient facing staff. Logistic regression was performed in R (12, 13).

### Severity of infection

To assess factors affecting severity of infection among seropositive staff we fit proportional odds logistic regression models in R using the MASS package (14). Severity classes were 1, 2, 3-4 (class 3 and 4 were grouped as only one member of staff reported severity 4). Age and sex were fit as predictor variables together, and with an interaction to understand the combined effect. The significance of the interaction was determined by likelihood ratio test of the two models. The predicted probabilities and their confidence intervals were visualised using the effects R package (15), (16). The assumption of proportional odds was assessed by refitting the model as two binary logistic regressions and comparing the estimated coefficients.

### Symptoms of seropositive and seronegative staff

To assess whether any of the reported symptoms discriminated seropositive from seronegative cases, data were collected on ten symptoms that have been associated with COVID19. Assessing all would require multiple testing and result in inflated false positive rates. To counteract this the p-value cut off for significance was adjusted following the Benjamini-Hochberg procedure based on the 10 tests (17). Furthermore, we tested the symptoms most likely to be discriminative first and stopped when the results were nonsignificant. Symptoms were ordered based on the difference in proportion of seropositive and seronegative staff reporting the symptom. Using logistic regression, the serostatus was predicted by the selected symptom. The significance of the symptom as a predictor was assessed by parametric bootstrapping using the R package pbkrtest (18).

The study was approved by Research Ethics Committee Wales, IRAS: 96194 12/WA/0148. Amendment 5 24.04.2020. All participants provided written, informed consent prior to enrolment in the study.

## Results

### Demographic of cohort

Results are reported for the first 500 of recruited staff members. Table 1a shows the demographic of the cohort by age, sex, ethnicity and work location in the cohort overall and by serostatus. The median age was 41 years and 3/5 of the staff recruited were women and 4/5 of white ethnicity. The majority of staff were patient facing (87%) and 25% worked in critical care (including a number of staff providing interhospital COVID19 patient transfers (data not shown)). Symptomatic staff screening commenced on week starting 6^th^ April 2020 and 10 of the staff recruited reported PCR positive. 2 additional staff were identified PCR positive as part of a point prevalence NHS screening initiative of asymptomatic staff. Recruitment occurred from 20^th^ April-10^th^ June 2020.

**Table 1a:** Cohort demographic for age, sex, ethnicity and work location and by serostatus by multiplex (N=493 classified). Values: N= (% of total). Age: years.*p=0.017: age of seropositive and seronegative cohort (MW-U).

|  |  | Total | Seropositive | Seronegative |
| --- | --- | --- | --- | --- |
| Number recruited |  | 500 | 70 (14) | 423 (86) |
| Age (yrs, median with IQR) |  | 42yr (33-51) | 40 (32-50) | 47yr (36-53)* |
| **Sex** | male | 133 (27) | 15 (11) | 118 (89) |
|  | female | 308 (62) | 42 (14) | 266 (86) |
|  | NA | 10 (10) |  |  |
| **Ethnicity group** | 1:white | 390 (78) | 45 (12) | 345 (88) |
|  | 2:mixed | 16 (3) | 4 (25) | 12 (75) |
|  | 3:asian | 79 (16) | 16 (20) | 63 (80) |
|  | 4:black | 11 (2) | 2 (18) | 9 (82) |
|  | 5: other | 1 (0.2) | 0 | 1 |
|  | NA | 3 (0.6) |  |  |
| **Work location** | Critical Care patient facing | 126 (25) | 10 (8) | 116 (92) |
|  | Non-Critical Care patient facing | 284 (57) | 40 (14) | 244 (86) |
|  | Non-patient facing | 63 (13) | 12 (19) | 51 (81) |
|  | NA | 27 (5) |  |  |

**Table 1b.**
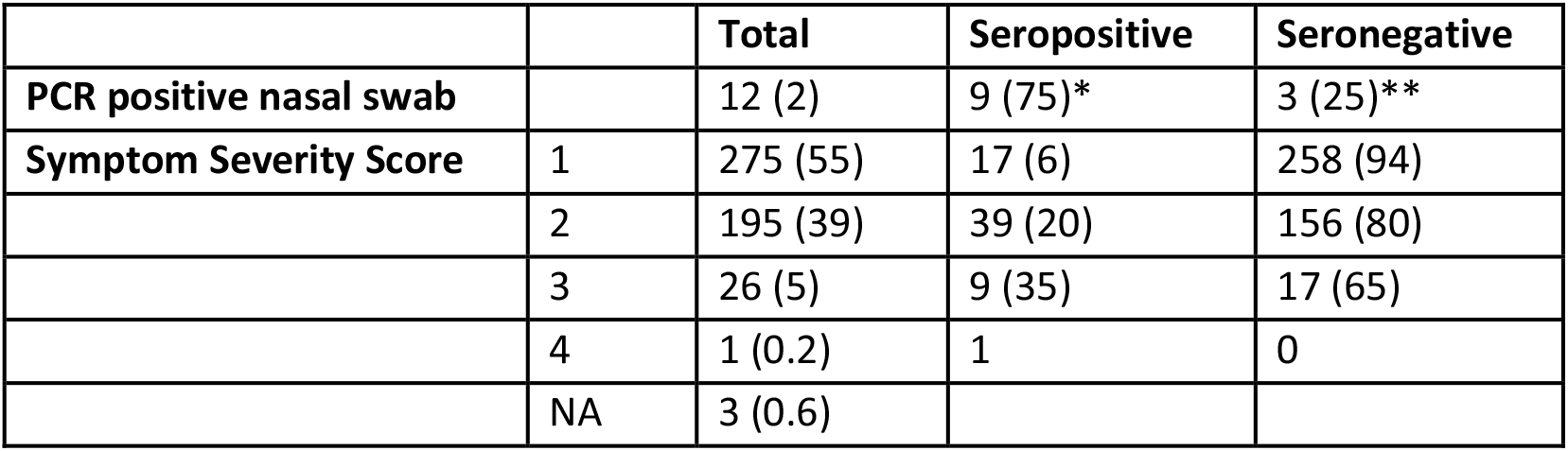
PcR Swab Positive and Symptom Severity Score by Serostatus. N= (% of total). *6 male,3 female; **3 female.

|  |  | **Total** | **Seropositive** | **Seronegative** |
| --- | --- | --- | --- | --- |
| **PCR positive nasal swab** |  | 12 (2) | 9 (75)* | 3 (25)** |
| **Symptom Severity Score** | 1 | 275 (55) | 17 (6) | 258 (94) |
|  | 2 | 195 (39) | 39 (20) | 156 (80) |
|  | 3 | 26 (5) | 9 (35) | 17 (65) |
|  | 4 | 1 (0.2) | 1 | 0 |
|  | NA | 3 (0.6) |  |  |

### Serostatus classification

Our SVM was able to successfully classify most samples in our training set. During cross validation the average sensitivity and specificity were 0.97 and 1.0 respectively. We were confident in our downstream analyses that seropositive samples are true positives due to the high specificity of our model (See Supplementary Figure 1a and 1b).

In total we classified 70 staff members seropositive and 423 seronegative based on S, RBD, and N binding. Seronegative individuals had low binding against all three antigens. However, some seropositive individuals had similarly low binding for some antigens. This overlap highlights the advantages of multi antigen screening. Several seropositive staff members had very low S and RBD binding but high N binding (Figure 1a and b). In contrast very few samples showed low N binding but high S or RBD binding. In fact, no sample in our training set had high RBD without similarly high N binding (Figure S1b). But in our staff member data we observed four samples with approximately zero N binding but elevated RBD binding (figure 1b). These samples were ostensibly seropositive but were classified as seronegative by our SVM because no similar samples were included in the training set. This highlights the diversity of serostatus and the importance of capturing that diversity when defining seropositivity and negativity.

**Figure 1.**
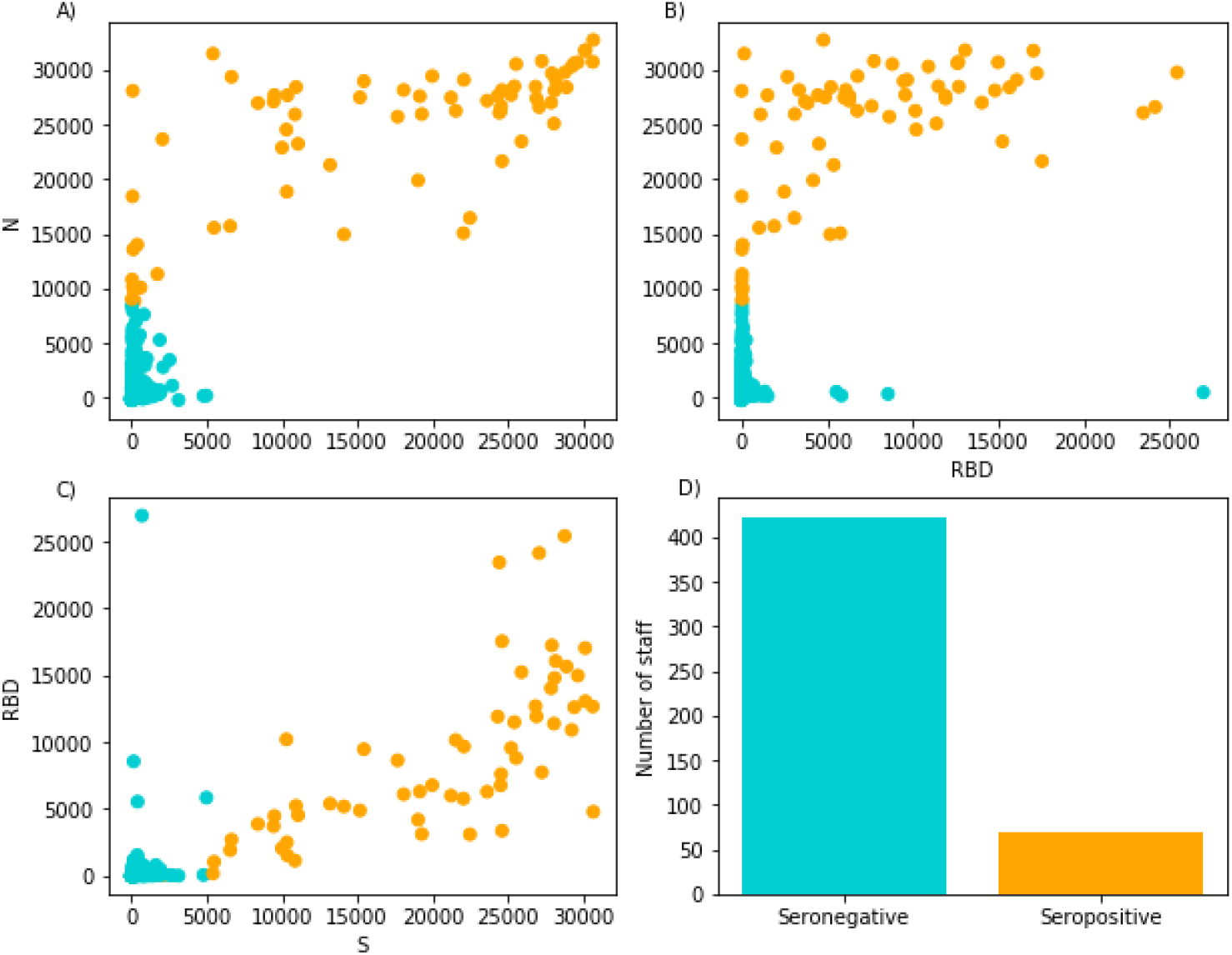
The relationship between A) N and S, B) S-RBD and N, C) S and S-RBD Luminex values (MFI) in Royal Papworth Hospital staff. D) shows the number of staff classified as seropositive or seronegative by the trained SVM. Serostatus is indicated by colour in all four panels.

**Figure 2.**
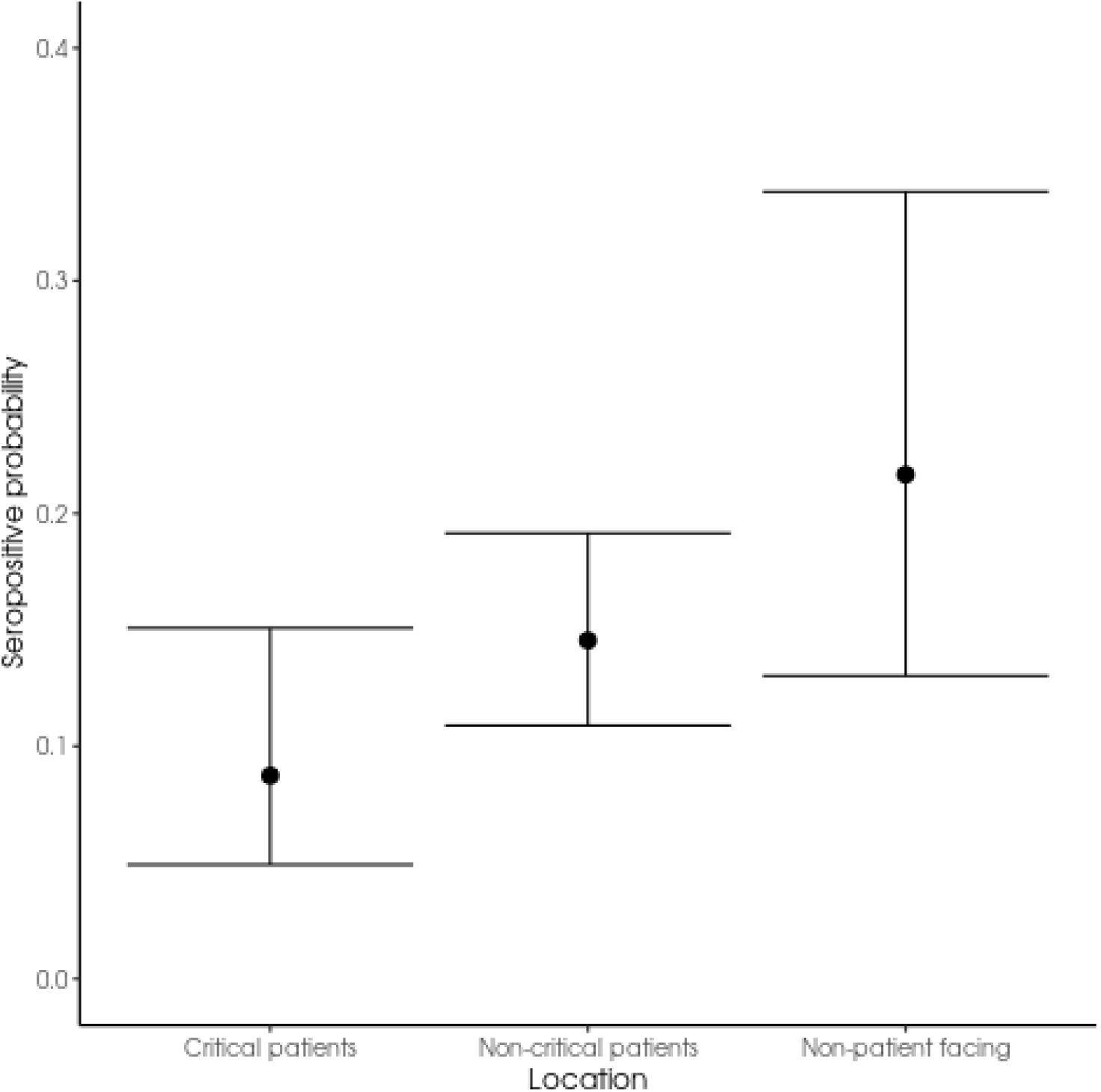
Probability of being classified as seropositive based on working location. Error bars show 95% confidence intervals.

**Figure 3.**
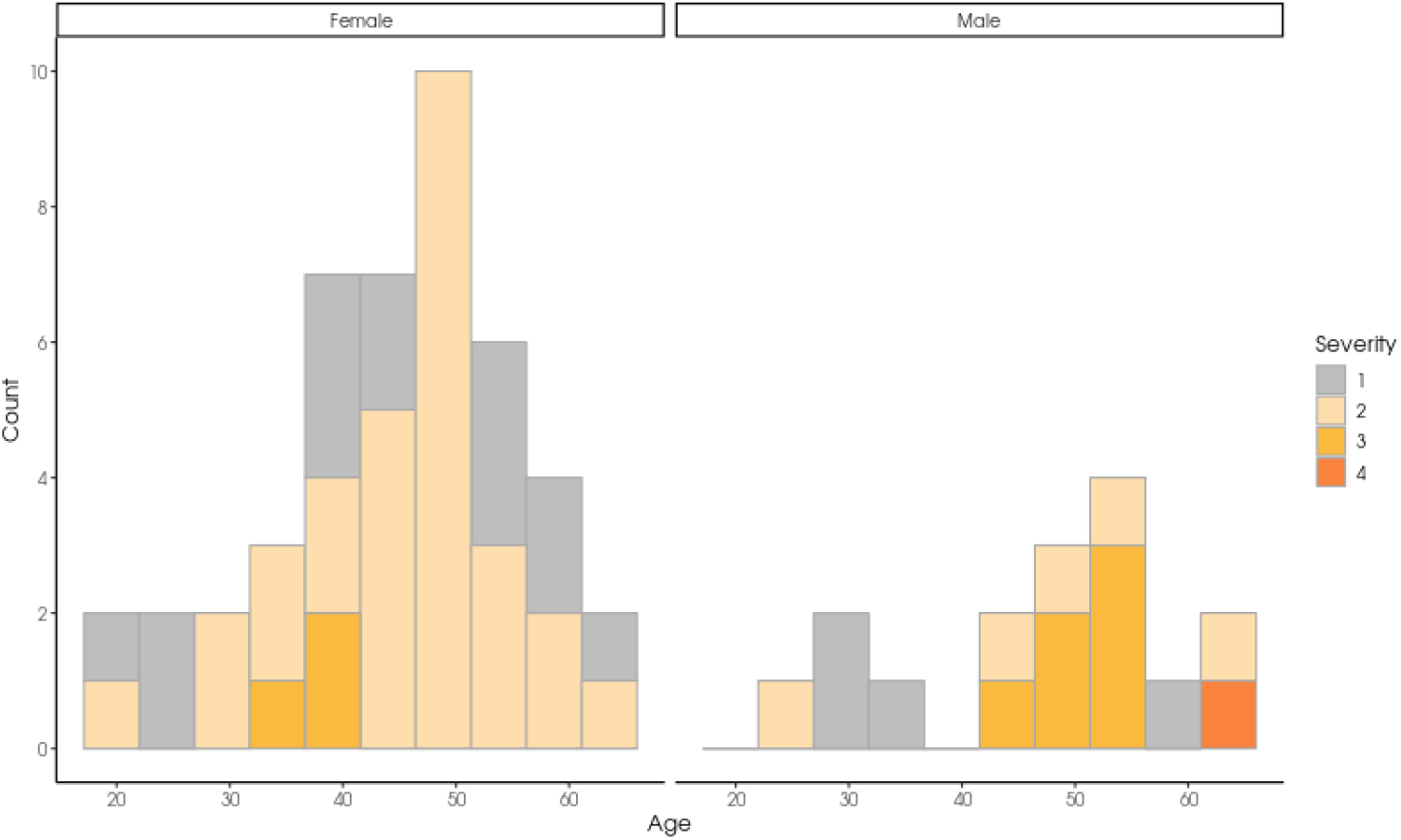
Stacked histogram of seropositive staff age split by sex and coloured by severity of infection.

**Figure 4.**
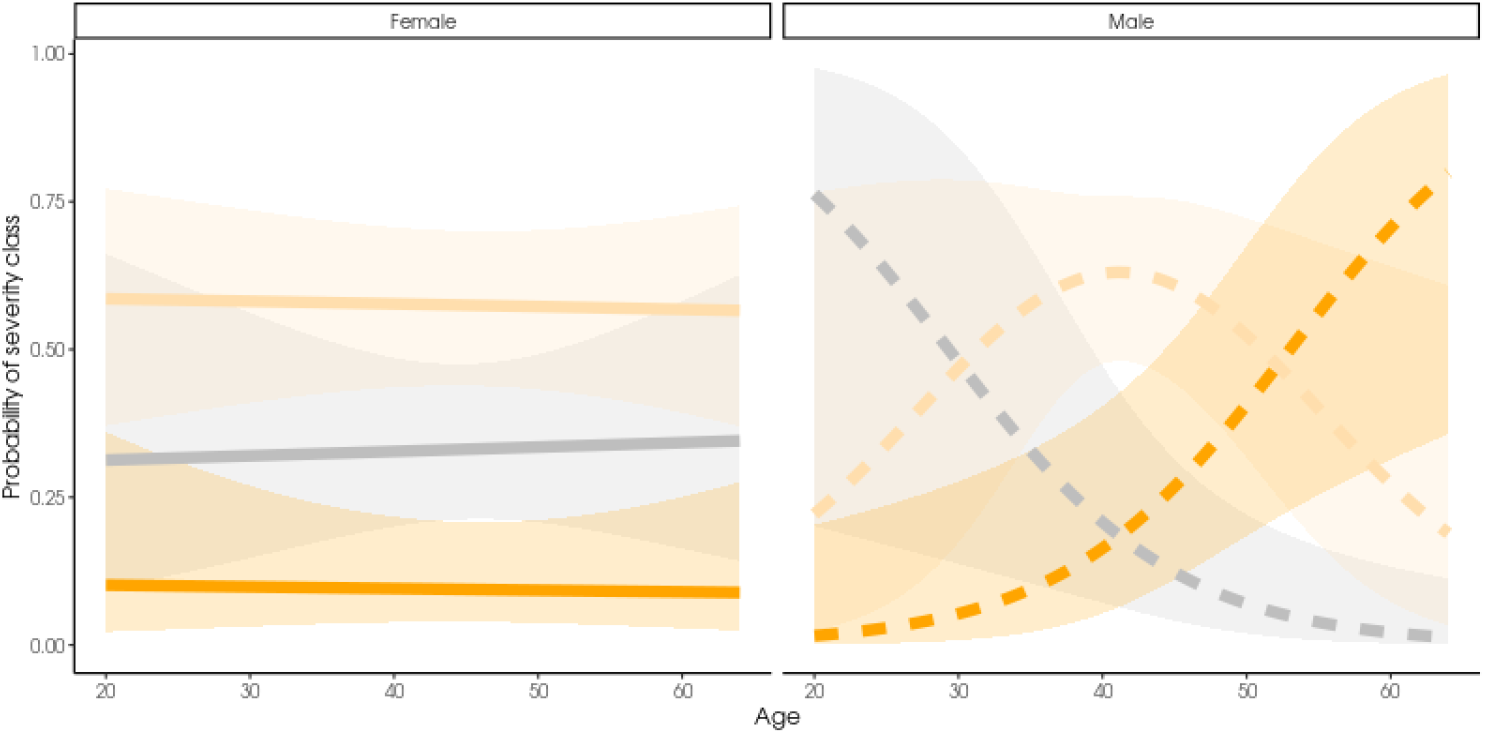
Probability of disease severity based on age and sex of seropositive staff. Severity of infection is indicated by colour; grey is class 1 (asymptomatic), pale orange is class 2, and dark orange is class 3 or 4 (grouped due to small sample size).

**Figure 5.**
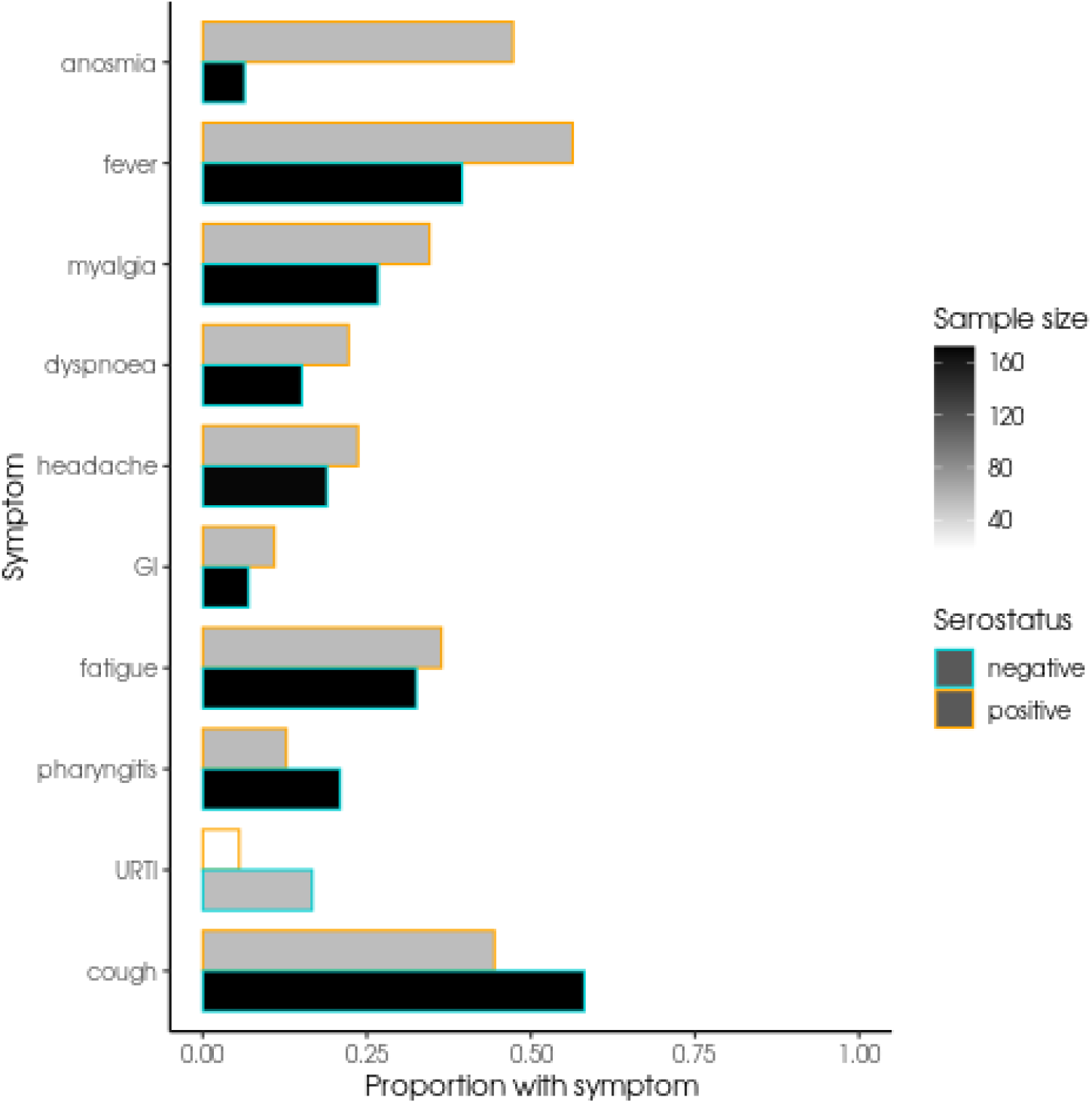
Proportion of seronegative and seropositive staff reporting specific symptoms. Bars outlined in orange indicate the proportion of seropositive staff with a symptom while bars outlined in turquoise indicate seronegative staff proportions. Symptoms are ordered by difference in proportion between seropositive and seronegative staff. The number of staff with symptom data available is indicated by the colour of the bar, darker bars indicated larger sample sizes.

### Serostatus by Demographic and Nature of Work

Serostatus by age, sex, ethnicity and work location is shown in Table 1a. A greater proportion of female HCW and those of non-white ethnicity were seropositive. Logistic regression was used to estimate the relationship between seropositivity and working location within Royal Papworth Hospital. 9% of staff in critical care were seropositive. Interestingly, staff working with critical care patients were significantly less likely to be seropositive than staff in non-patient facing roles (β=-1.06, SE=0.44, Z=-2.39, p=0.017). Serostatus for non-critical care patient facing staff was not significantly different compared to non-patient facing staff (β=-0.49, SE=0.36, Z=-1.14, p=0.172) at 15 and 22% seropositive respectively.

### Symptom reporting, COVID19 clinical severity score and Serostatus

A high proportion of staff (45%) reported possible COVID19 symptoms in the 4 months prior to recruitment of whom 5% reported symptoms consistent with moderate/severe disease (Table 1b). The majority of these staff were found to be seronegative, however the relative proportion of seropositive staff increased with symptom score. Of the few staff (N=12, 6 male and 6 female) who had PCR swab results from time of symptoms (N=10) or through staff surveillance screening (N=2), 3 PCR positive staff (all female) were seronegative, 2 of whom reported a history of symptoms consistent with moderate severity COVID19 (one was asymptomatic). There was no difference in month of symptom reporting and serostatus at time of recruitment (data not shown).

Analysis of symptom severity in the seropositive cohort indicated that infection severity depended on age according to a sex specific relationship (sample size of 45 women, 16 men). Male staff’s risk of severe infection rose steeply with age but age had little effect on the risk of severe infection among female staff (age β=-0.00, SE=0.03, sex_male_ β=-4.53, age:sex_male_ β=0.13). The intercept for asymptomatic cases was –0.85 (SE=1.25) and for severity class 2 the intercept was 2.11 (SE=1.30). A likelihood ratio test found this interaction to significantly improve model fit (−2LL = 5.7, p = 0.017). There was no relationship between age or sex and symptom severity for seronegative staff.

The effect of age appears to be driven by male cases as when a model was fit to only female staff an age effect was not significant (−2LL = 0.02, p = 0.89).

### Symptoms of seropositive and seronegative staff

The only symptoms which could significantly predict serostatus were anosmia and fever. Anosmia β=2.57, SE=0.41, p<0.001, fever β=0.68, SE=0.31, p=0.031, myalgia was also tested but was non-significant β=0.37, SE=0.33, p=0.301. However, after accounting for multiple testing only anosmia remained a significant predictor of serostatus.

## Discussion

We performed multiplex testing to evaluate antibody binding to N, S, and RBD proteins. Our study suggests that detection of N protein specific IgG is the most sensitive method for seroprevalence screening. A high proportion of seropositives also demonstrated both S and RBD IgG responses, and there was good correlation in serostatus across the 3 antigens. Whilst the value of N protein only based platforms has generated some debate in view of the weaker correlation with viral neutralising activity compared with S and RBD binding assays (19), our data supports other reports showing that N protein based assays are most sensitive for detection of serum responses(20, 21). Within the seropositive cohort, we found no relationship between symptom score and the spectrum of binding activity to the 3 antigens tested (data not shown). Multiplex platforms offer simultaneous, high through put screening for multiple antigens using small serum samples and as such provide an attractive platform for large scale serum analysis of responses to multiple antigens. Excellent sensitivities and specificities are further increased by its multiplexed nature, which may be in particular useful in earlier stages as some patients may not yet have fully seroconverted and may only have anti-N antibodies (22-25). Correlation between ELISA and multiplex is generally strong (26) and this represents an attractive platform for clinicians and researchers in characterising acute and convalescent serum responses an establishing serological correlates of protective immunity.

We demonstrate that staff working with COVID-19 patients in a critical care environment have lower prevalence of serum antibodies to SARS-CoV-2 than those working in patient facing non critical care, and that non-patient facing staff have the highest seroprevalance of the three cohorts assessed. It is unclear if differences in seroprevalence between staff groups represents risk of workplace infection. However at the time of the study, most non-clinical facing staff worked in open plan offices where the wearing of face masks was not policy at that stage and social distancing was inconsistently adhered to. In contrast, staff in critical care adhered to guidance strictly. These results may provide some re-assurance to critical care staff and staff running hospital transfers including ECMO implementation, that the current PPE guidelines are fit for purpose. A potential confounder to this interpretation is that the participants in the study were self-selected and as such this may introduce bias however these results complement those of other published studies, that staff working in the critical care environment are at lower risk of workplace acquired infection compared with other health care environments (27). Whether all hospital workers at increased risk of COVID19 is unclear noting that demographic matching is required to answer this question. At the time of staff sampling, regional seroprevalence was reported to be approximately 8% (28).

Nearly 50% of the staff recruited reported a history of symptoms that they felt may have represented SARS-CoV-2 infection in the preceding 1-4 months, however 14% of staff were found to be seropositive. With considerable overlap between symptom reporting in early COVID-19 and that of other common respiratory viruses, identifying symptoms that are more specific to SARS-CoV-2 infection is important, not least as screening resources even in the most wealthy countries are limited with the aim of targeting screening at the individuals presenting with symptoms most likely representing COVID19 infection. Analysis of free field symptom reporting, and correcting for multiple tests, we found that anosmia was the most discriminating symptom for seropositive status in our cohort. This is consistent with recently published reports of COVID-19 confirmed patients reporting anosmia additional to fever and continuous cough as being one of the most discriminating symptoms of COVID-19 (29) (30, 31). Recent onset anosmia has now been added to the 3 symptoms used to diagnose COVID-19 and prioritise track a trace screening in the UK (32). As we move into winter when prevalence of infection with other respiratory viruses will soar, ongoing evaluation of the reliability of using these three symptoms (anosmia, persistent cough and fever) to direct COVID-19 testing will be important noting that in our cohort, 25% of seropositive staff reported no history of symptoms that they considered consistent with possible COVID-19. These results complement existing data generated through both seroprevalence and virus surveillance in health care workers and the population, demonstrating that 15-40% of individuals have asymptomatic infection in the UK with highest prevalence in younger cohort (27, 31) (33)

More detailed interpretation of these data is limited by the low numbers of staff with contemporaneous PCR swab results. Of the 12 staff who reported previous SARS-CoV-2 PCR positive by nasal swabbing (10 symptomatic, 2 though surveillance), 3 were seronegative (all female and 2 moderately symptomatic at time of swabbing). Serum samples for all staff were taken at least 4 weeks after symptom reporting so this does not reflect premature serum sampling rather it suggests seroprevalence may under-represent infection rates. Diagnostic thresholds were established using acute phase sera from patients with moderate to severe disease and as such may have reduced sensitivity for detecting history of exposure particularly in those with asymptomatic or mild infection in whom short lived or no seroconversion has been reported (34). In addition, some exposed individuals have been reported to have evidence of cell mediated immune responses in the absence of serum antibodies(12). How prevalent this is not yet known.

COVID-19 Symptom severity scoring showed a striking relationship between age and symptom severity in seropositive male staff. There was no relationship between age and symptom severity in female seropositive staff or seronegative staff. Being of male sex and aged >40 years are reported a risk factors for severe COVID19 (12). Older males are at increased risk of pneumonic complications from other viruses such as influenza (35) and other causes of community acquired pneumonia (36) however on multivariate analysis the male sex as an independent predictor of disease severity is often not sustained. Detailed analysis of the relationship between age and sex in seasonal and pandemic influenza shows that sex hormones and virus specific pathogenesis of disease influence the relationship between age, sex and disease severity in man and murine models of disease (37). Understanding the differences between the sexes in pathogenesis (and possibly immunopathogenesis, including antibody mediated exacerbation of disease) of COVID-19 will be important to inform targeted disease prevention and treatment strategies.

Whether individuals who do not seroconvert are at increased risk of re-infection compared with those who develop high serum antibody titres remains an open question. This unknown generates considerable anxiety for staff who – despite widely published concerns relating to an ‘immunity passport’ -are generally self-re-assured that if they develop antibodies post infection that they may be less at risk of subsequent re-infection.

This study has a number of limitations. Most significantly, that nasal swabbing for SARS-CoV-2 PCR tests were not available for symptomatic staff early in the pandemic when most of our staff reported symptoms that they felt may have represented COVID-19. Some care should be taken when interpreting the exact relationship between severity of infection and age of men as relatively few male staff members were classified as seropositive. However, many other studies have similarly reported increased incidence of severe COVID-19 in men especially older men (38, 39) (40)

Large cohort, longitudinal studies with paired swab and serum samples additional to symptom reporting are now running. In the UK, the Sarscov2 Immunity & REinfection EvaluatioN longitudinal health care worker surveillance study, SIREN (41) where swab and serum samples are collected at 2-4 weekly intervals in large cohorts, in addition to symptom reporting, will provide the power to define in detail the relationship between serum response, symptom severity and re-infection risk in HCW by demographic. Although it is important to acknowledge that many staff identified as being at increased risk of severe COVID-19 have been shielding and/or working remotely and may be under-represented in these workplace based cohort studies.

In conclusion, this data presented suggest: 1. staff working in critical care environments looking after large numbers of COVID-19 patients and those transferring acutely unwell patients for escalation of care, are not at increased risk of COVID-19 infection compared with staff in non-clinical roles. 2. Anosmia most reliably predicts seropositive infection in this cohort. 3. Serum antibody status likely significantly under-represents infection prevalence. 4. Further investigation is needed to understand the relationship between serum antibody response and disease severity by demographics noting the role of different antibodies in developing COVID-19 disease remains unclear.

## Data Availability

Analysis code will be made available on an online repository on publication.

## Acknowledgements

Funding: Royal Papworth Hospital NHS Trust: R&D pump priming. This work is now funded by the UKRI and NIHR MC_PC_20016: HICC: Humoral Immune Correlates for COVID19: Defining protective responses and critical readouts for Clinical Trials of Vaccines and Therapeutics. We would like to thank staff from RPH recruited to the study, Dr Ian Smith^1^, Professor William Schwaeble and Dr Javier Castillo-Olivares Pallardo^2^ for critical review of the manuscript and Leo Kiss of MRC LMB for helping with SARS-CoV-2 N protein reagents.

