## Supplemental Figure 1 for "Critical care workers have lower seroprevalence of SARS-CoV-2 IgG compared with non-patient facing staff in first wave of COVID19"

Supplementary File

Figure 1:
S1a. ROC curves established with training data set from PCR confirmed SARS-CoV2 positive patients and pre-pandemic negative control samples for individual N, S and RBD antigens.
S1b. The relationship between A) N and S, B) N and RBD C) S and RBD Luminex values (MFI) in COVID-19 patients.

S1a

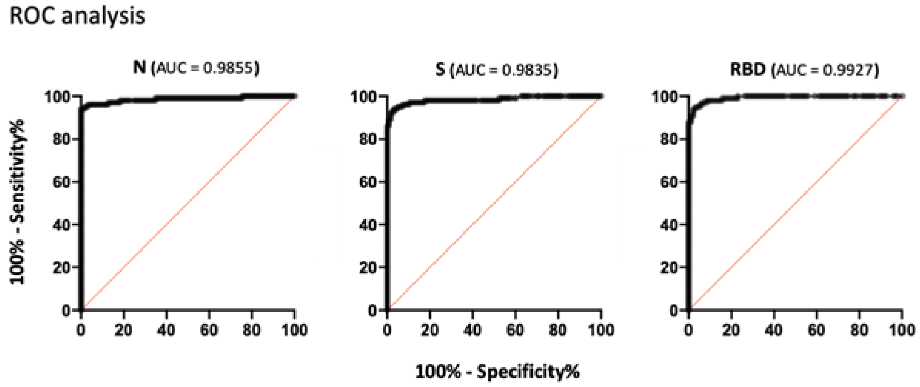


S1b

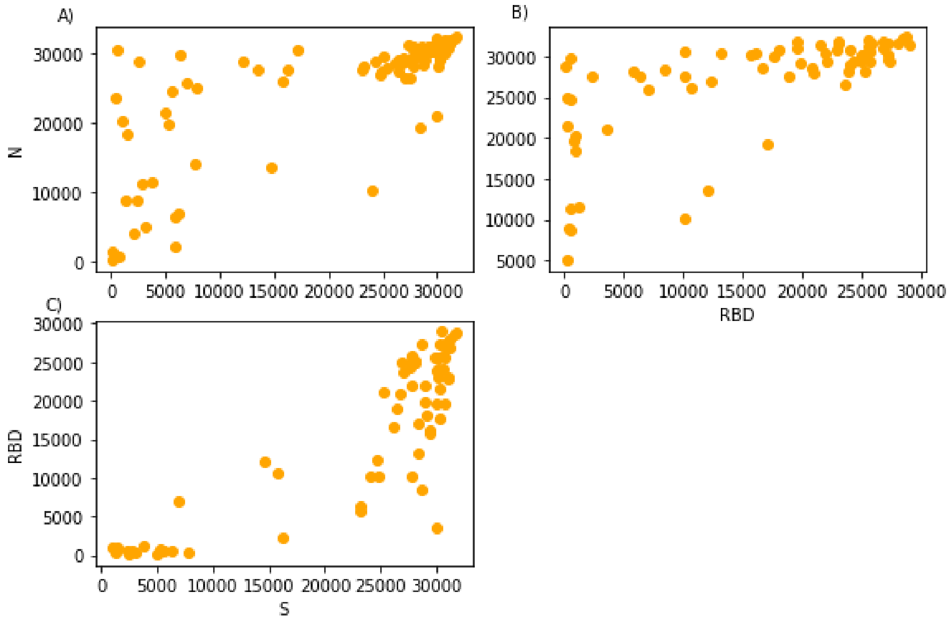
